# The Long-Term Impacts of Dementia on Preventive Care Utilization and Health Behaviors

**DOI:** 10.1101/2023.10.24.23297477

**Authors:** Zhuoer Lin, Xi Chen

## Abstract

Dementia has large impact on individuals’ decision making, independent living, and wellbeing. Identifying early signals of dementia risk may offer people more time to prepare for the future, helping to delay the onset or slow the progression of dementia. Using the 1995-2018 waves of Health and Retirement Study, we offer novel evidence on the impacts of dementia on a rich set of preventive care utilization and health behaviors. Leveraging both within- and between-individual variations in an event study design, we characterize long-term dynamic changes in preventive care and health behaviors relative to the incidence of dementia and find early behavioral indicators of the disorder. We show that relative to the group of people who never develop dementia during the study periods, people with dementia have consistent and escalating declines in the use of cholesterol test, dental visit, prostate test and mammogram around the incidence of dementia. Significant declines are also found in physical activities and social engagement. Importantly, we demonstrate that the behavioral changes can occur up to 6 years before the incidence of dementia; and these patterns are absent in other chronic or acute conditions. The results are robust to sample selection, model specification, and the further control of aging and cohort effects. Overall, our findings highlight the salient impact of dementia risk on preventive care utilization and health behaviors, which may increase individuals’ vulnerability to health shocks. Detecting early signals of dementia and facilitating targeted interventions are thus called for to prevent individuals from adverse behavioral and health consequences.

## 1. Introduction

Dementia, a highly prevalent neurological disorder, has far-reaching effects on various aspects of individuals’ lives, including decision-making, independent living, and overall wellbeing. It imposes immense burdens not only on affected individuals but also on their families and society as a whole. Presently, there are approximately 55 million people worldwide living with dementia, a number projected to increase substantially in the coming decades. The financial impact of dementia is also substantial, with estimated costs exceeding 1.3 trillion USD annually (Alzheimer’s Association, 2023; Alzheimer’s Disease International, 2020).

Given the profound consequences of dementia, it is crucial to take proactive measures during the early stages to enhance individuals’ resilience against brain pathologies and mitigate the adverse effects of neurological disorders (Alzheimer’s Association, 2021). By identifying earlier signals of dementia risk, we can provide individuals with valuable time to prepare for the future, thereby aiding in delaying the onset or slowing the progression of dementia.

Emerging evidence indicates that risky financial decisions and financial errors often serve as early indicators of dementia (Marson, 2001; Nicholas et al., 2021; Widera et al., 2011). A recent study further demonstrates that signs of adverse financial events may occur years before a formal diagnosis of dementia, which holds informative values for dementia early detection (Nicholas et al., 2021). However, there remains a dearth of evidence documenting the impacts of dementia on health care utilization and health behaviors (Kang and Xiang, 2020; Lin et al., 2016), limiting our understanding of early presentation of the disorder.

The impact of dementia on health-related behaviors can be consequential. Deficits in memory and other cognitive domains create substantial cognitive barriers that hinder individuals from accessing health care and engaging in preventive behaviors (Dickson et al., 2007; Feil et al., 2012; Lovell et al., 2019). Additionally, the stigma associated with dementia often leads to heightened reluctance among individuals to seek care (Alzheimer’s Disease International, 2019). These behavioral changes, in turn, increases individuals’ vulnerability to dementia risks and health shocks, exacerbating the deterioration of cognitive function.

However, existing research on the behavioral changes in dementia remains partial and limited. Studies on health behaviors are often cross-sectional (Hall et al., 2006) or fail to adequately account for changes in cognitive status (Anstey et al., 2009; Daly et al., 2015; Hall et al., 2006; Kang and Xiang, 2020). Similarly, research on healthcare utilization frequently suffer from small sample size (Eaker, 2002; Hajduk et al., 2013; Ton et al., 2017) or focuses primarily on inpatient care utilization and costs, overlooking preventive care utilization (Desai et al., 2019; Lin et al., 2016; Lugo-Palacios and Gannon, 2017; Ton et al., 2017). Moreover, studies on health care utilization and health behaviors rarely account for the temporal relationship between cognitive and behavioral changes (Mehta et al., 2010; Yang et al., 2021), limiting the interpretability of the results (Angrisani and Lee, 2019). Importantly, no study to date has documented the long-term dynamic changes in health-related behaviors relative to the incidence of dementia (Fereshtehnejad et al., 2018; Nicholas et al., 2021), making it challenging to understand the early evolution of the behavioral changes.

Using U.S. nationally representative longitudinal data from the Health and Retirement Study (HRS) spanning up to 24 years of follow-up (1995-2018), we provide one of the first quantifications of the impacts of dementia on a comprehensive set of preventive care utilization and health behaviors. Particularly, we characterize the dynamic changes in preventive care and health behaviors relative to the incidence of dementia, revealing early behavioral indicators of the disorder. Specifically, based on a biannual dementia status ascertainment, we identify individuals who have ever developed dementia during the study periods; and for each observations around the incidence of dementia, we use propensity score matching to select a 1:1 matched control group with similar characteristics who have never developed dementia. Employing an event study design, we quantify changes in behavioral outcomes between the dementia sample and the matched control sample before and after the incidence of dementia.

Our results indicate that incidental dementia is associated with significant reductions in preventive care utilization, including cholesterol tests, dental visits, mammograms, and prostate tests. We also observe significant declines in physical activities and social engagement. Moreover, these effects tend to increase over time, with more pronounced impacts occurring in the years following the onset of dementia compared to incidental changes. Importantly, these behavioral changes can manifest years before the incidence of dementia. For instance, the use of preventive care begins to decline approximately 4-6 years before the incidence, indicating early presentation of the disorder. The magnitude of the decline is also substantial, ranging from 5 to 20 percentage points (pp) around dementia incidence. These findings are robust to various alternative approaches to sample selection and model specifications, as well as more flexible control for age and cohort effects.

Furthermore, the observed patterns specific to dementia are not observed for other health shocks, such as the incidence of hypertension, diabetes, hip fracture, and myocardial infarction, highlighting the unique impact of dementia. Overall, our results underscore the significant influence of dementia risk on preventive care utilization and health behaviors, as well as the early presentation of the disorder.

This study contributes to the literature in three key aspects. First, it provides initial evidence on the impacts of dementia on a comprehensive set of health-related behaviors, many of which have not been thoroughly examined in previous research. Second, compared to studies using medical claims data, our use of survey-based cognitive assessments enables more accurate capturing of the transition of cognitive status and reduces the likelihood of underdiagnosis issues related to dementia. Finally, employing a dynamic event study design, this study characterizes the long-term changes in health-related behaviors both before and after the onset of dementia, revealing important early behavioral indicators of the disorder.

The remainder of the paper is organized as follows. Section 2 presents the data and variables used in the study. In Section 3, we outline the study design and identification strategy. The main results are presented in Section 4. Section 5 discusses the results of robustness analyses and placebo tests, and Section 6 concludes.

## 2. Data and Variables

### 2.1 Data Source

The data for this study are obtained from the Health and Retirement Study (HRS), which is a nationally representative longitudinal study of adults aged 50 or older in the United States. The HRS survey is conducted approximately every two years from 1995 to 2018, and each wave includes interviews with around 19,000 participants (Power et al., 2021). The HRS Core Interview collects comprehensive information on demographics, socioeconomic conditions, health, cognition, health care utilization, and other relevant factors. Starting in 2004, the HRS added a lifestyle questionnaire to gather additional data on psychosocial factors such as subjective well-being and social relationships (Sonnega et al., 2014).

For this study, the variables are mainly constructed based on the RAND HRS Longitudinal File, which is a cleaned and harmonized dataset that enhances the comparability of variables across survey waves (RAND Center for the Study of Aging, 2022). Some variables that are not available in the RAND HRS are constructed using HRS public data files from 1995 to 2018.

### 2.2 Preventive Care Utilization and Health Behaviors

Two sets of outcome variables are investigated in this study: preventive care utilization and health behaviors. For preventive care, self-reported information is collected in each wave regarding whether individuals had a memory diagnosis, a blood test for cholesterol, a dental visit, a mammogram (for females) to screen for breast cancer, and a PSA blood test (for males) to screen for prostate cancer in the past two years. Participants report if they have ever been told by a doctor that they have memory-related diseases, including Alzheimer’s disease and related dementias. This measure allows us to track if individuals are aware of or get diagnosis of their memory problems relative to the incidence of dementia. Dental visits are measured in every wave for all participants. Cholesterol tests, mammograms, and prostate tests are collected every other wave for all participants, and prior wave values are carried forward to ensure continuity in measurements (RAND Center for the Study of Aging, 2022). Mammograms are asked among female participants, while prostate tests are asked among male participants. Additionally, a dichotomous measure is constructed to indicate whether individuals had any of the regular preventive care, including dental visits, cholesterol tests, mammograms (for females) or prostate tests (for males) in the past two years, providing an overall pattern of preventive care utilization.

While some preventive care measures, such as mammograms and prostate tests, may only apply to certain age groups according to clinical guidelines (Howard et al., 2013; Qin et al., 2017), other measures, such as dental visits and cholesterol tests, generally apply to all groups. Involving all these preventive care measures may complement each other in strengthening our investigation into the intrinsic link between dementia and preventive care use dynamics.

As for health behaviors, two key modifiable factors for dementia are considered: physical activities and social engagement (Livingston et al., 2020). These factors are applicable to the entire population and have clear implications for individuals’ well-being. In each wave since 2004, the frequencies of vigorous activities (e.g., running, swimming), moderate activities (e.g., gardening, stretching exercises), and light activities (e.g., vacuuming, laundry) are consistently measured. For each type of physical activity, a dichotomous variable is constructed to indicate whether individuals participate in the activities more than once a week (1 = physically active) or not (0 = physically inactive). Regarding social engagement, a summary score ranging from 0 to 4 is constructed to assess the extent of individuals’ engagement in four social behaviors: participation in any social groups, clubs, or organizations at least once a month; monthly contact with children, relatives, and friends (Crowe et al., 2021; Steptoe et al., 2013).

### 2.3 Dementia Assessment

In the HRS, cognition is assessed using the Telephone Interview for Cognitive Status (TICS), which yields a score ranging from 0 to 27. The TICS comprises three cognitive tests: immediate and delayed word recall (0-20 points) to assess memory, a serial sevens subtraction test (0-5 points) to assess working memory, and a backward counting test (0-2 points) to evaluate speed of mental processing. If participants are unable to complete these cognitive tests independently, their cognition is evaluated based on data from proxy respondent interviews. This evaluation involves a total cognitive scale that incorporates proxy assessment of the respondent’s cognition, instrumental activities of daily living (IADLs), and the interviewer’s assessment of the respondent’s cognition (Crimmins et al., 2011; Langa et al., 2008).

Using an established algorithm, individuals in each wave are classified as “demented” if their 27-point cognitive scores fall below 7 (0-6 points), or their proxy-respondent cognitive scales fall into “demented” category (Crimmins et al., 2011; Langa et al., 2008). As dementia status can be determined in each wave from 1995 to 2018, with up to 24 years of follow-up, it allows us to identify the potential transition from a “pre-dementia” state to a state of dementia over time.

### 2.4 Covariates

In addition to behavioral outcomes and dementia status, a rich set of covariates are considered in this study. These covariates include demographic factors (age, sex, race/ethnicity, marital status, number of children), socioeconomic status (education, wealth level), health insurance coverage (Medicare enrollment, Medicaid enrollment, military health plan enrollment, private or employer-based health insurance coverage, long-term care insurance coverage), and health status and comorbidities (functional limitations, depressive symptoms, hypertension, diabetes, cancer, lung diseases, heart diseases, psychiatric problems, arthritis). Most of the covariates are time-varying, with the exceptions being sex, education, and race/ethnicity.

## 3. Study Design and Identification Strategy

### 3.1 Sample Selection and Propensity Score Matching

Two analytical samples are included in the study to examine the differential trends in behavioral outcomes resulting from dementia. As shown in Figure 1, the first group consists of individuals in the dementia sample who have experienced the transition from a “pre-dementia” stage to a “dementia” stage during the study period (1995-2018). Only participants with at least one consecutive wave of data before the onset of dementia, with no reversion back to a non-dementia stage afterwards, are included. All person-waves before and after the incidence of dementia are considered for analysis.

**Figure 1.**
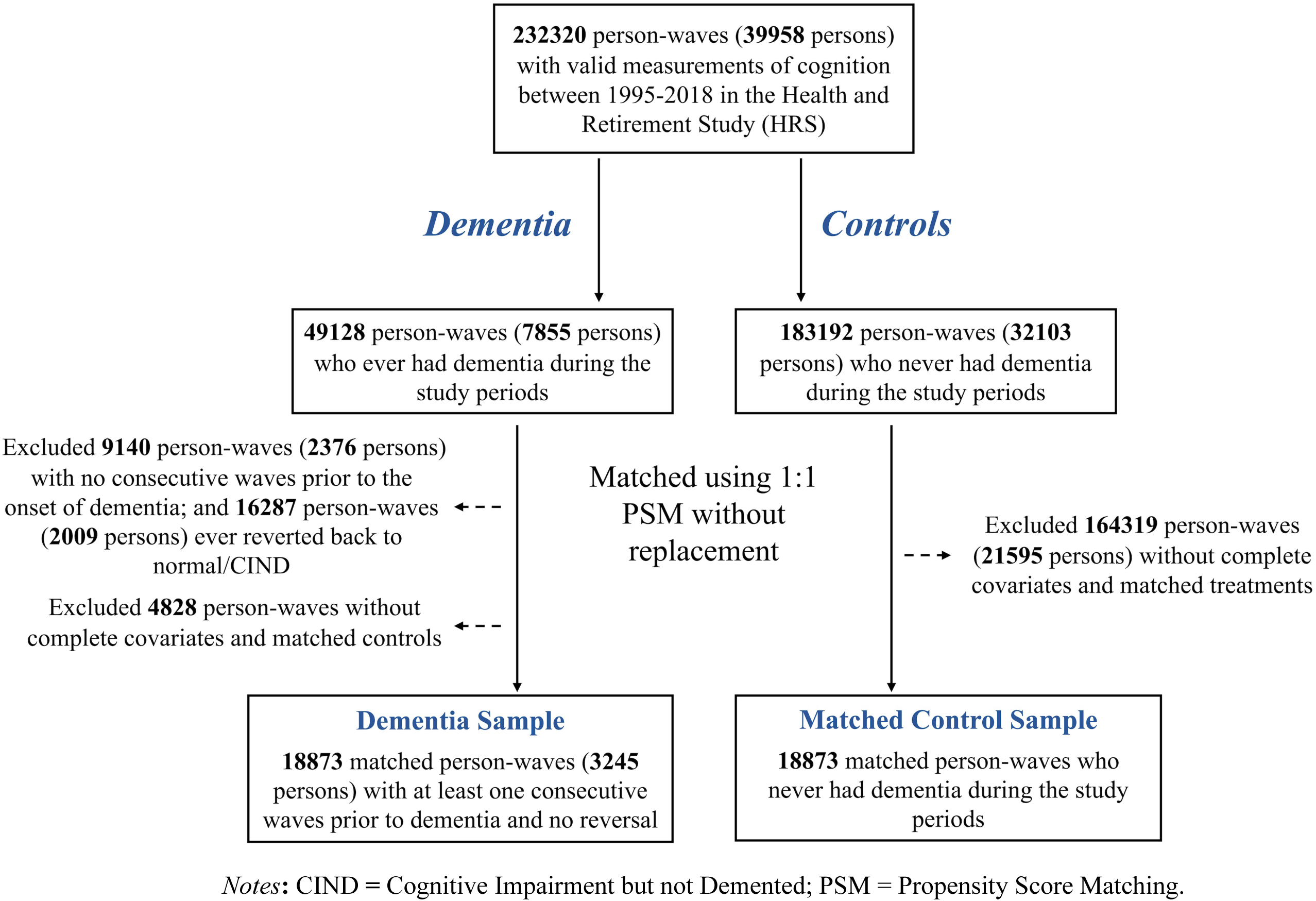
Flow chart of sample selection.

For the control sample, which comprises individuals who never develop dementia during the study period, propensity score matching (PSM) is employed to identify 1-to-1 matched controls. A logistic regression model is used to estimate propensity scores, with treatment assignment (dementia vs. control) as the dependent variable and the aforementioned covariates as independent variables. The propensity scores accordingly capture the predicted probability of developing dementia during the study period. Based on these estimated scores, observations from the dementia sample and the control sample are matched 1-to-1 without replacement, using a caliper of 0.20 standard deviations of the propensity scores (Austin, 2010; Desai et al., 2019; Lunt, 2014). Figure 2 presents the balance of covariates between the dementia sample and the control sample before and after matching. After matching, the standardized % biases substantially reduce, with biases falling within the 10% threshold, indicating strong balance across the covariates. Overall, the PSM results in 18,873 matched pairs of dementia and control sample with similar characteristics.

**Figure 2.**
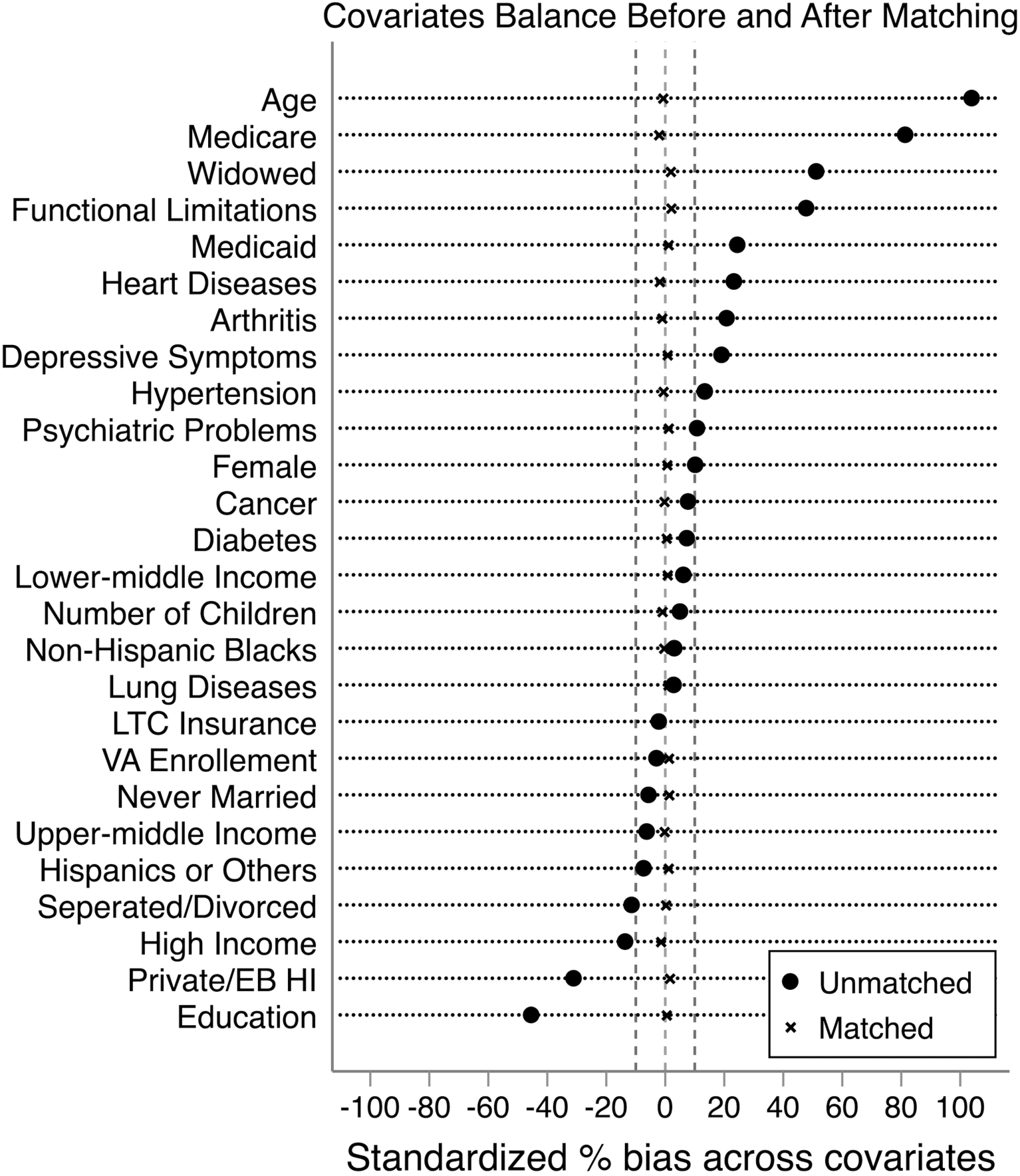
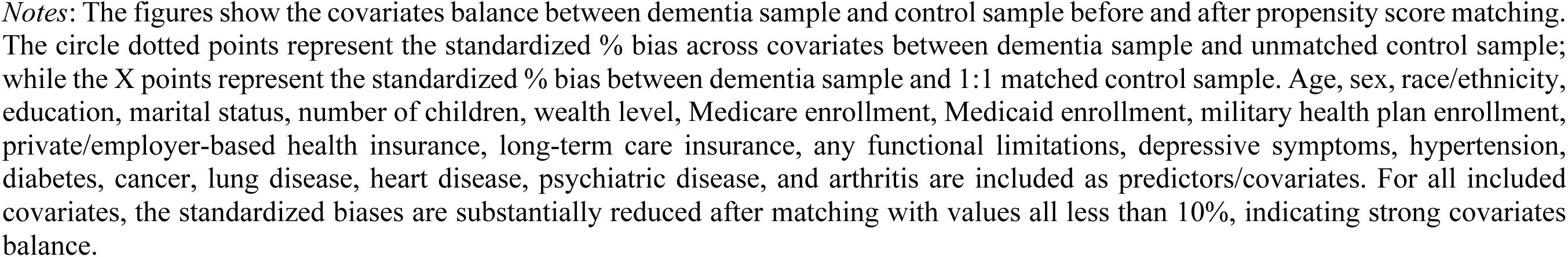
Covariates Balance Before and After Propensity Score Matching.

### 3.2 Event Study Design

To examine the long-term impact of dementia on preventive care utilization and health behaviors, an event study design is employed to analyze the dynamic changes in behavioral outcomes before and after the incidence of dementia (Angrisani and Lee, 2019; Nicholas et al., 2021). This design leverages both within-individual and between-individual variations and compares behavioral outcomes between the treatment group (individuals who eventually develop dementia) and the matched control group (individuals who never develop dementia during the observation period).

Time indicators (dummies) are included in the model to estimate the differences in behavioral outcomes between the treatment and control groups at each time point relative to the incidence of dementia. The model is specficied as follows:

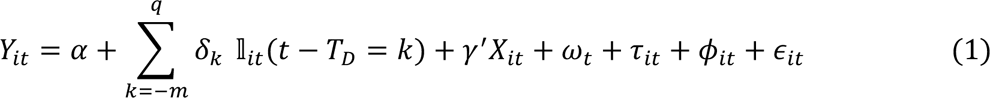

where the dependent variable *Y*_*it*_ represents whether individual *i* use specific preventive care or engage in certain health behaviors as measured in year *t*. The time indicators 𝕝_*it*_(*t* − *T*_*D*_ = *k*) capture the relative time from the first occurance of dementia (*T*_*D*_) for individual *i* who develop dementia during the study periods. The refernece group consists of matched individuals who never develop dementia, allowing for differentiation between changes induced by cognitive disorder and normal aging patterns (Nicholas et al., 2021). The analysis includes up to 8 time periods (16 years) prior to the first occurrence of dementia and up to 3 time periods (6 years) following the onset of the disorder. The post-dementia period is relatively short due to potential dropouts or lack of interviews after developing dementia. *X*_*it*_ represents a comprehensive set of observed time-invariant and time-varying covariates included to control for individual heterogeneity. Additionally, years (*ω*_*t*_), regions (*τ*_*it*_), and year-by-regions (*ϕ*_*it*_) fixed effects are included to control for the overall time trend, geographical differences, and region-specific time trend (Qian et al., 2021). The coefficients *δ*_*k*_ captures the changes in behavioral outcomes at each time period *k* for the dementia sample relative to the matched control sample.

The estimation is conducted using a linear probability model rather than a Logit or Probit model. This choice allows for a direct interpretation of the coefficients, whereas Logit/Probit models may yield inconsistent estimates when a large number of indicator variables are included (Freedman, 2008; Gomila, 2021).

## 4. Main Results

### 4.1 Sample Characteristics

Table 1 presents the characteristics of the sample. The dementia and control samples have similar demographics, socioeconomic status, health insurance coverage, health status, and comorbidities. On average, 63% of the sample is female, and 74% are non-Hispanic White. The average age is 75 years old, and the average years of education is 12.

**Table 1.**
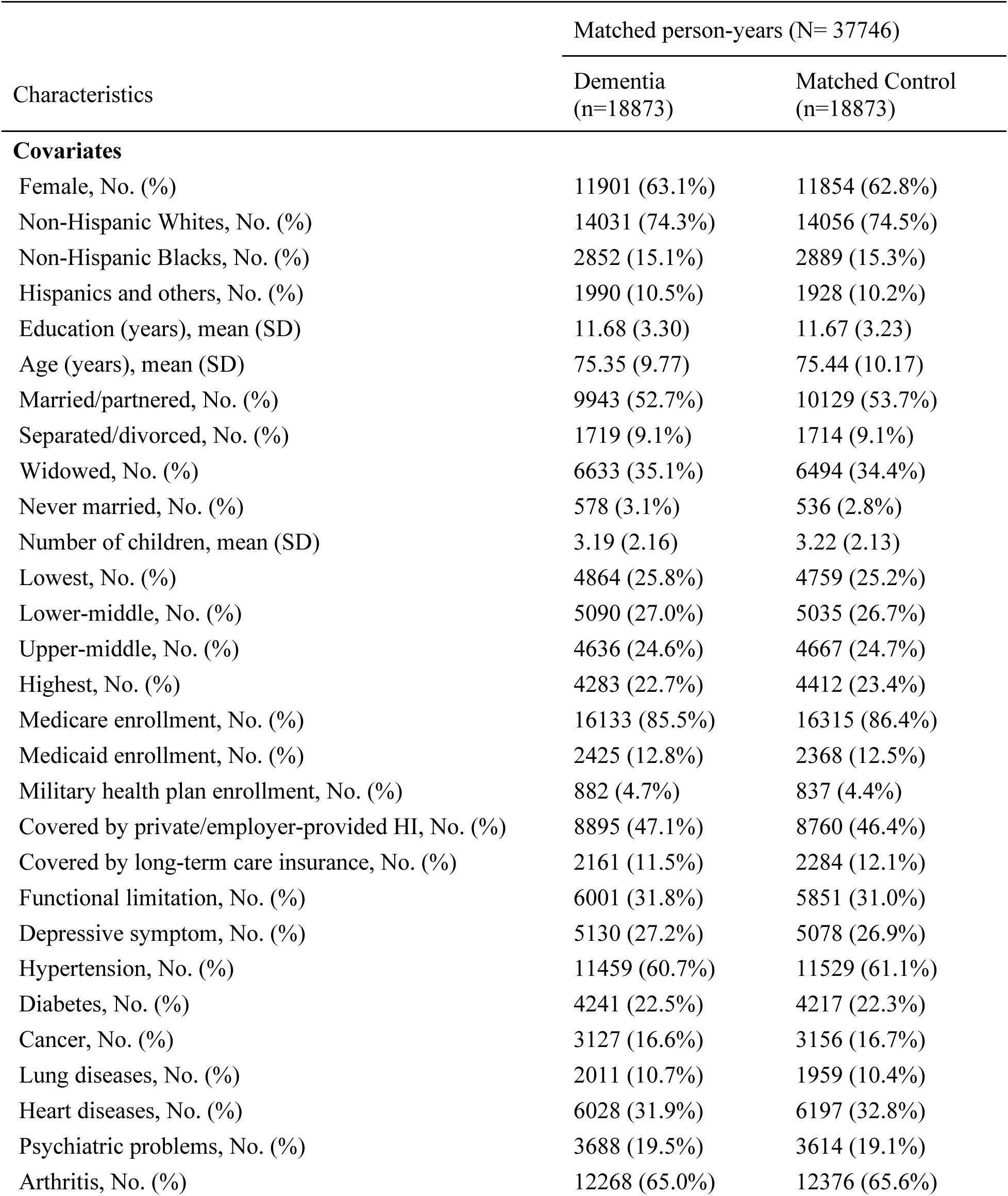
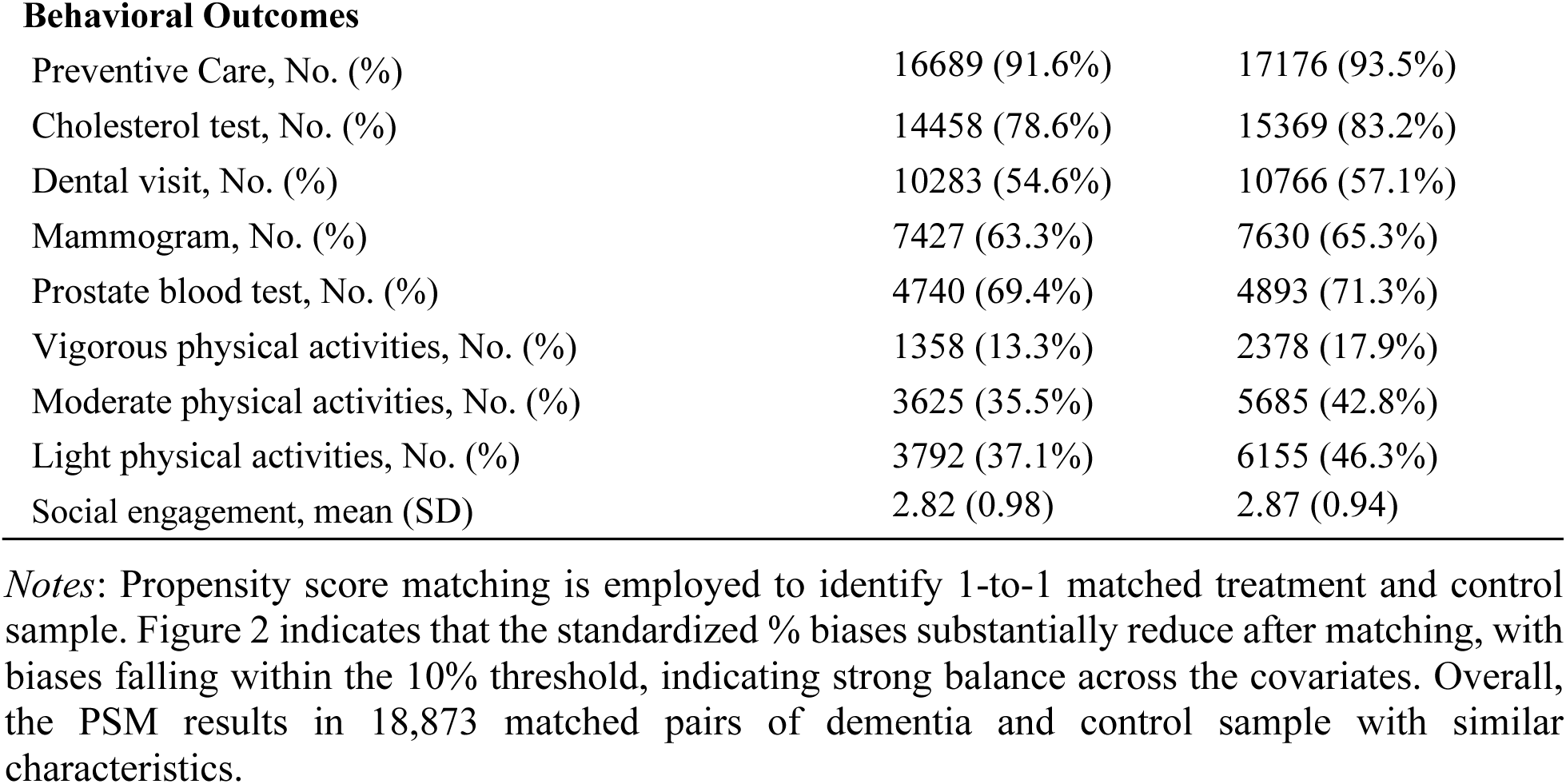
Characteristics of matched sample by dementia status.

| Characteristics | Matched person-years (N= 37746) |  |
| --- | --- | --- |
|  | Dementia<br>(n=18873) | Matched Control<br>(n=18873) |
| **Covariates** |  |  |
| Female, No. (%) | 11901 (63.1%) | 11854 (62.8%) |
| Non-Hispanic Whites, No. (%) | 14031 (74.3%) | 14056 (74.5%) |
| Non-Hispanic Blacks, No. (%) | 2852 (15.1%) | 2889 (15.3%) |
| Hispanics and others, No. (%) | 1990 (10.5%) | 1928 (10.2%) |
| Education (years), mean (SD) | 11.68 (3.30) | 11.67 (3.23) |
| Age (years), mean (SD) | 75.35 (9.77) | 75.44 (10.17) |
| Married/partnered, No. (%) | 9943 (52.7%) | 10129 (53.7%) |
| Separated/divorced, No. (%) | 1719 (9.1%) | 1714 (9.1%) |
| Widowed, No. (%) | 6633 (35.1%) | 6494 (34.4%) |
| Never married, No. (%) | 578 (3.1%) | 536 (2.8%) |
| Number of children, mean (SD) | 3.19 (2.16) | 3.22 (2.13) |
| Lowest, No. (%) | 4864 (25.8%) | 4759 (25.2%) |
| Lower-middle, No. (%) | 5090 (27.0%) | 5035 (26.7%) |
| Upper-middle, No. (%) | 4636 (24.6%) | 4667 (24.7%) |
| Highest, No. (%) | 4283 (22.7%) | 4412 (23.4%) |
| Medicare enrollment, No. (%) | 16133 (85.5%) | 16315 (86.4%) |
| Medicaid enrollment, No. (%) | 2425 (12.8%) | 2368 (12.5%) |
| Military health plan enrollment, No. (%) | 882 (4.7%) | 837 (4.4%) |
| Covered by private/employer-provided HI, No. (%) | 8895 (47.1%) | 8760 (46.4%) |
| Covered by long-term care insurance, No. (%) | 2161 (11.5%) | 2284 (12.1%) |
| Functional limitation, No. (%) | 6001 (31.8%) | 5851 (31.0%) |
| Depressive symptom, No. (%) | 5130 (27.2%) | 5078 (26.9%) |
| Hypertension, No. (%) | 11459 (60.7%) | 11529 (61.1%) |
| Diabetes, No. (%) | 4241 (22.5%) | 4217 (22.3%) |
| Cancer, No. (%) | 3127 (16.6%) | 3156 (16.7%) |
| Lung diseases, No. (%) | 2011 (10.7%) | 1959 (10.4%) |
| Heart diseases, No. (%) | 6028 (31.9%) | 6197 (32.8%) |
| Psychiatric problems, No. (%) | 3688 (19.5%) | 3614 (19.1%) |
| Arthritis, No. (%) | 12268 (65.0%) | 12376 (65.6%) |
| **Behavioral Outcomes** |  |  |
| Preventive Care, No. (%) | 16689 (91.6%) | 17176 (93.5%) |
| Cholesterol test, No. (%) | 14458 (78.6%) | 15369 (83.2%) |
| Dental visit, No. (%) | 10283 (54.6%) | 10766 (57.1%) |
| Mammogram, No. (%) | 7427 (63.3%) | 7630 (65.3%) |
| Prostate blood test, No. (%) | 4740 (69.4%) | 4893 (71.3%) |
| Vigorous physical activities, No. (%) | 1358 (13.3%) | 2378 (17.9%) |
| Moderate physical activities, No. (%) | 3625 (35.5%) | 5685 (42.8%) |
| Light physical activities, No. (%) | 3792 (37.1%) | 6155 (46.3%) |
| Social engagement, mean (SD) | 2.82 (0.98) | 2.87 (0.94) |

*Notes:* Propensity score matching is employed to identify 1-to-1 matched treatment and control sample. Figure 2 indicates that the standardized % biases substantially reduce after matching, with biases falling within the 10% threshold, indicating strong balance across the covariates. Overall, the PSM results in 18,873 matched pairs of dementia and control sample with similar characteristics.

Averaging across the study period, there are notable differences in behavioral outcomes between the dementia and control samples. The dementia sample has a lower proportion of individuals utilizing any preventive care, cholesterol test, dental visit, mammogram, and prostate test compared to the matched control sample. Additionally, the dementia sample is less likely to engage in vigorous, moderate, and light physical activities, as well as social engagement, compared to the control sample.

### 4.2 Differential Trends in Behavioral Outcomes between Dementia and Control Sample

Figure 3 illustrates the temporal trends in preventive care utilization for both the dementia and matched control samples around the incidence of dementia without regression adjustment. Regarding memory-related diagnoses, a marked rise is observed around the time of dementia incidence. However, it is important to note that a substantial portion of the dementia sample remains without timely diagnosis. Over 60% of the sample is unaware of their memory problems at the time of dementia incidence. Even 4-6 years following the incidence, approximately 30% of the sample still lacks diagnosis. In contrast, memory-related diagnoses for the control group show negligible changes.

**Figure 3.**
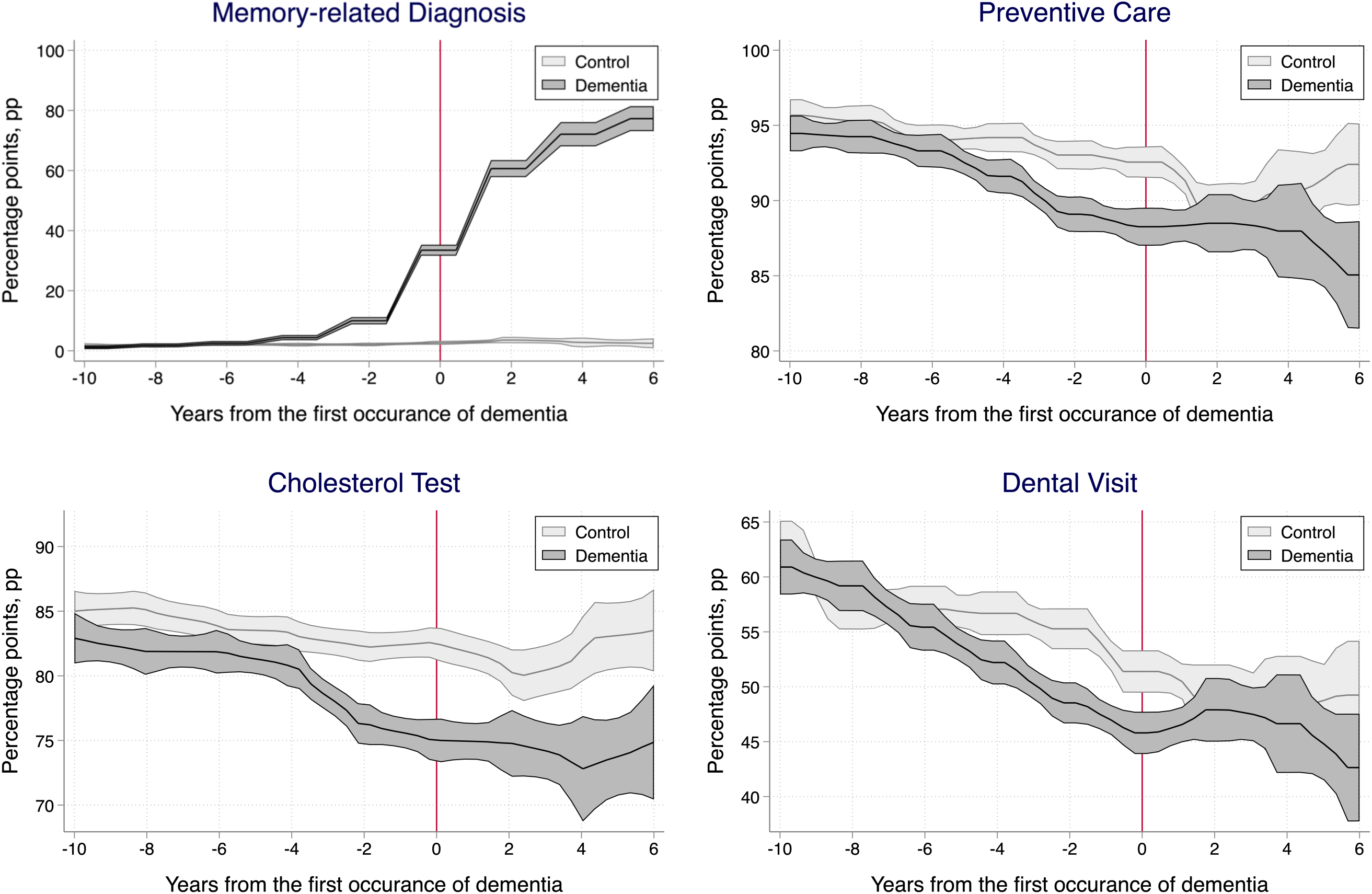
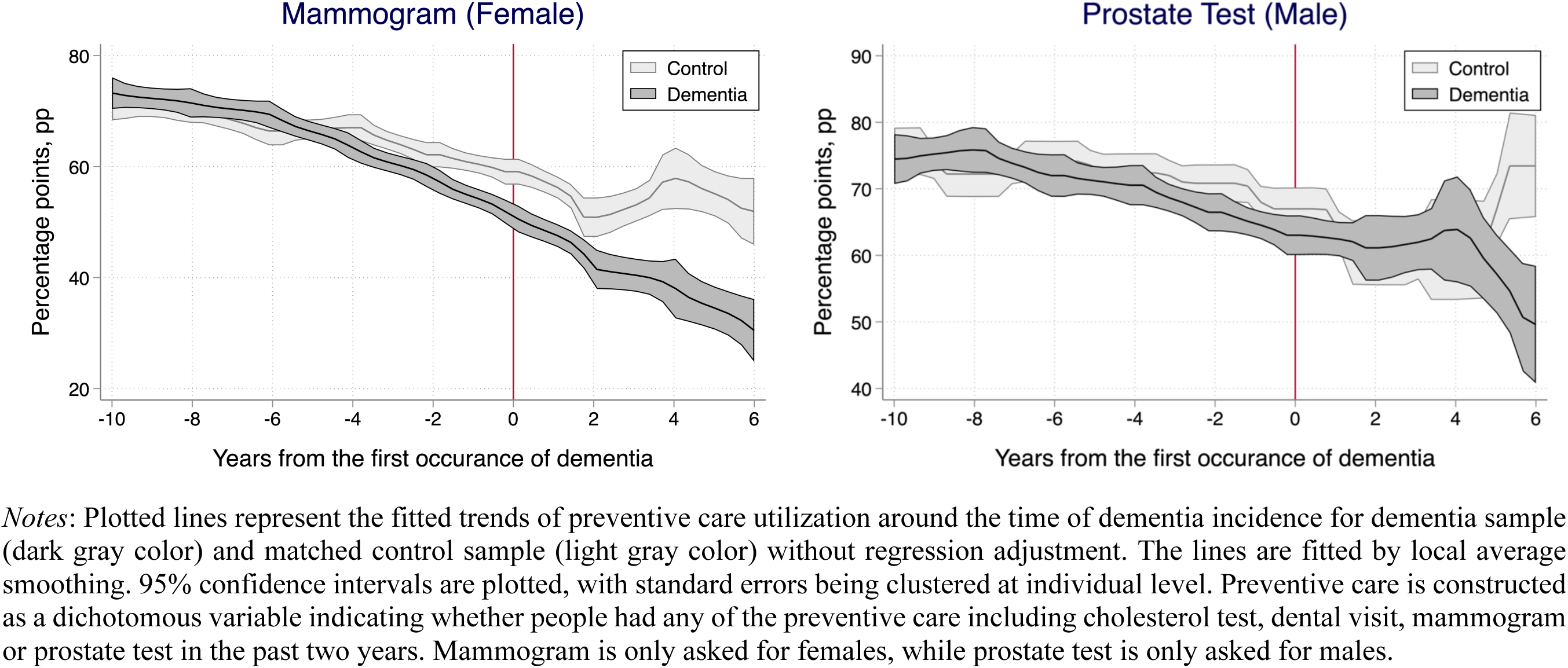
Trends in preventive care utilization before and after dementia for dementia sample and matched control sample during 1995-2018 without regression adjustment.

Conversely, the patterns differ for other preventive care measures unrelated to memory-related diseases. Approximately 6-10 years prior to the incidence of dementia, both the dementia and control samples exhibit comparable levels and trends in preventive care utilization. However, a distinct pattern emerges around 4-6 years prior to dementia, with the dementia sample experiencing a noticeable decline in preventive care utilization compared to the control sample. As time progresses, the gap between the two groups widens further. Similar diverging trajectories are observed for cholesterol tests, dental visits, mammograms, and prostate tests. Notably, the declines in cholesterol tests and dental visits are particularly pronounced during earlier time periods.

For healthy behaviors, the differential trends between the dementia and control samples are even more pronounced, as shown in Figure 4. The dementia sample experienced significant reductions in physical activities and social engagement beginning 2-4 years prior to the incidence of dementia. Notably, the gaps between the dementia and control samples could reach up to 20 percentage points for physical activities by the time of dementia incidence.

**Figure 4.**
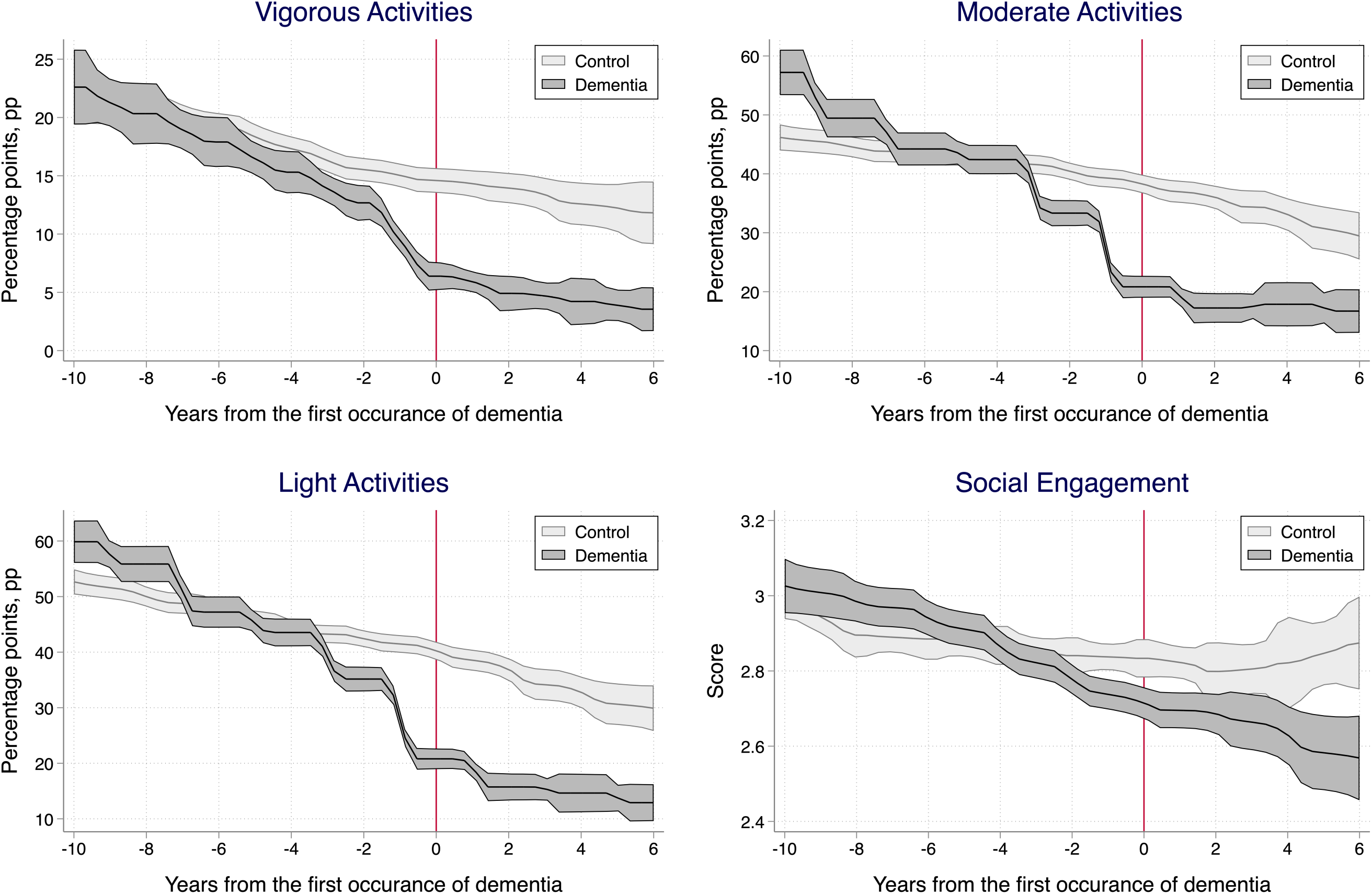
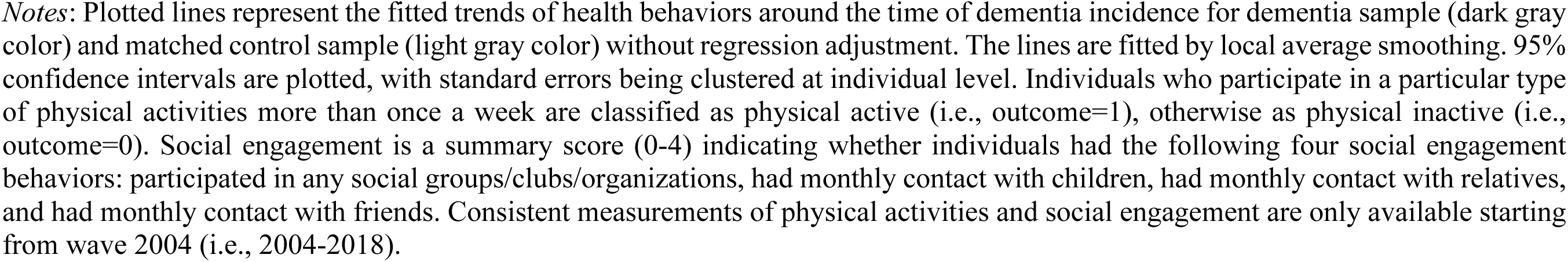
Trends in health behaviors before and after dementia for dementia sample and matched control sample during 2004-2018 without regression adjustment.

### 4.3 Event Study Estimates: The Long-Term Impact of Dementia on Behavioral Outcomes

Figures 5 and 6 present the event study estimates, which demonstrate the differences in behavioral outcomes between the dementia and matched control samples at each time point relative to the incidence of dementia, after adjusting for individual and geographic heterogeneity as well as common trends.

**Figure 5.**
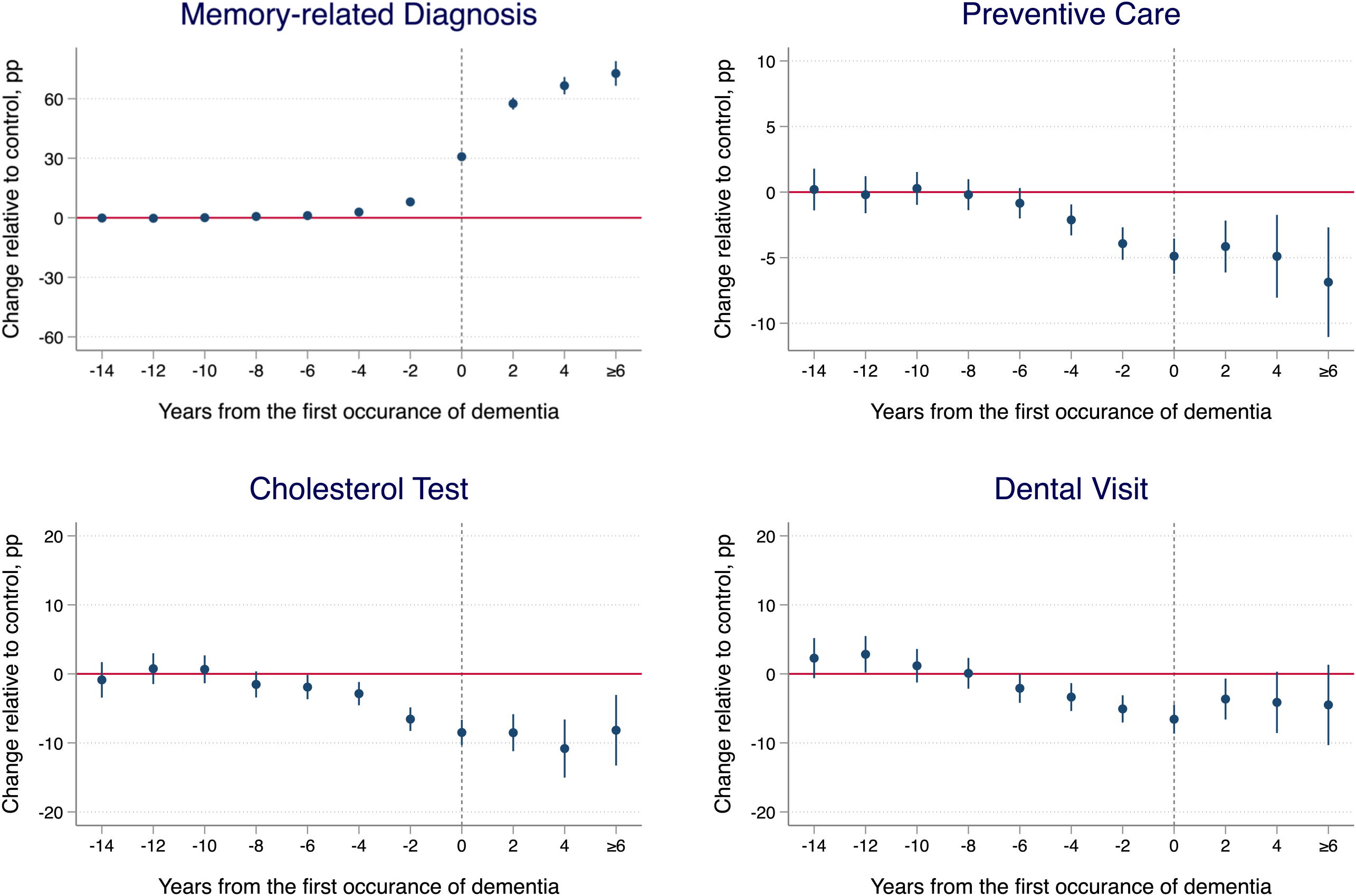
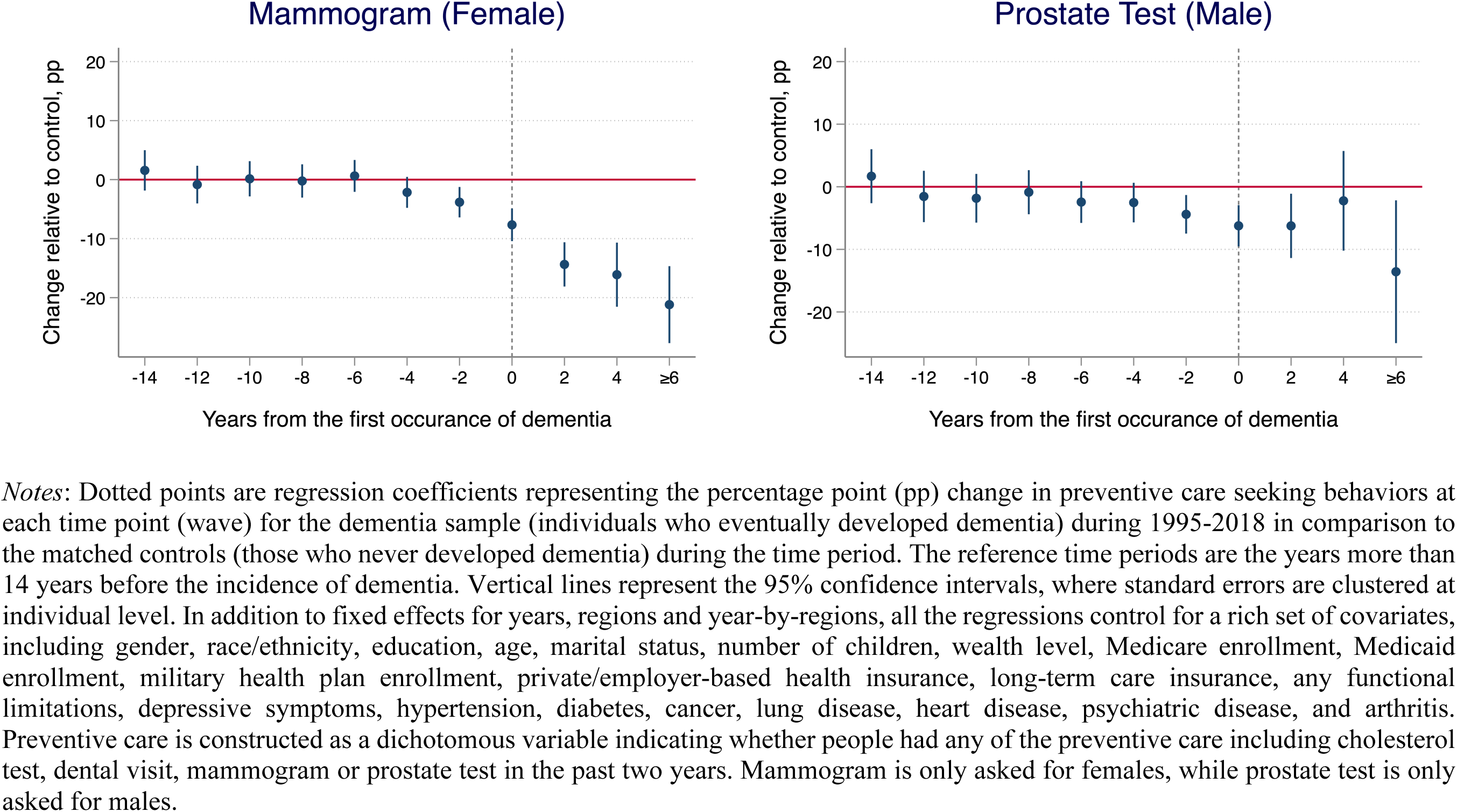
Change in preventive care utilization before and after dementia for the dementia sample relative to the matched control sample during 1995-2018.

**Figure 6.**
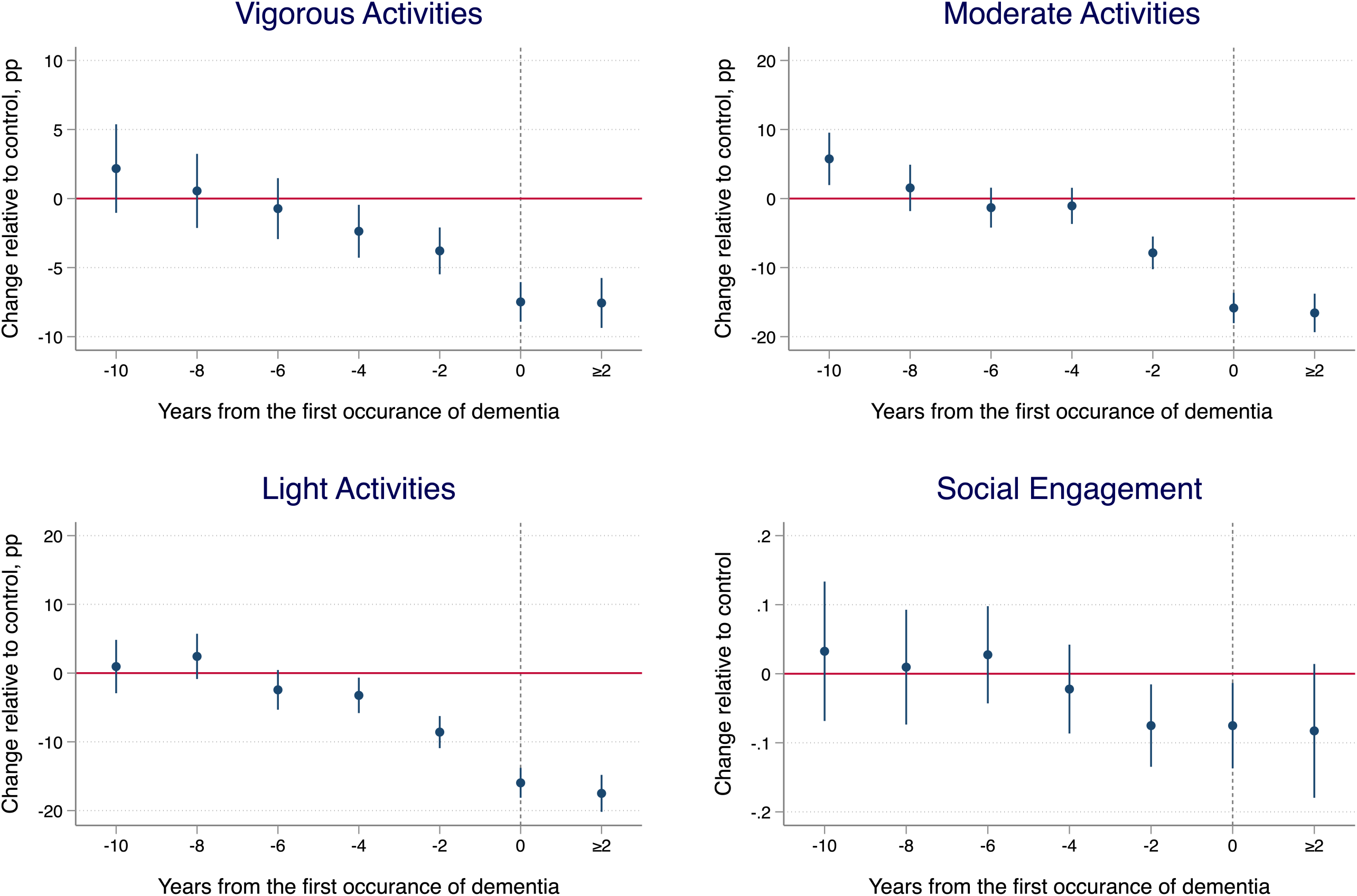
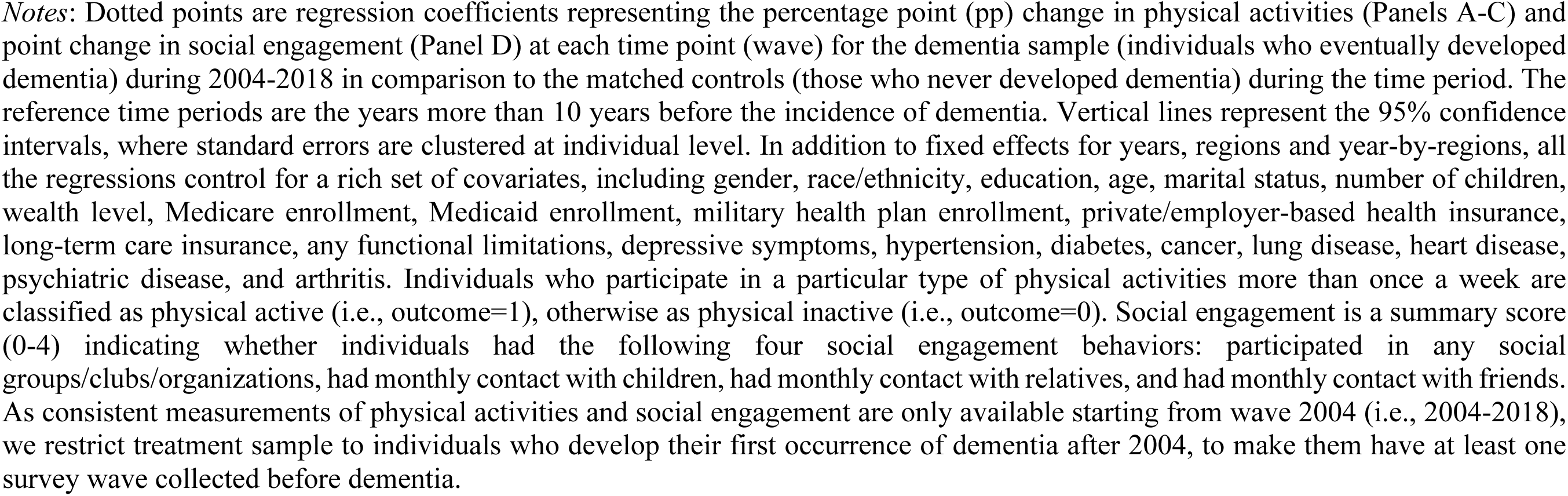
Change in health behaviors before and after dementia for the dementia sample relative to the matched control sample during 2004-2018.

Consistent with the unadjusted patterns, we witness a rise in memory-related diagnoses for individuals in the dementia sample compared to the control sample, while there is a substantial decline in other preventive care measures unrelated to memory-related diseases. Importantly, our results reveal significant differences in preventive care utilization between the dementia and control samples that emerge years before the incidence of dementia (Figure 5). Compared to the matched control sample, individuals who develop dementia exhibit significantly lower preventive care utilization starting 4 years prior to dementia incidence. Moreover, the reductions in preventive care utilization tend to increase notably over time, and these patterns are evident across different types of preventive care, including cholesterol test, dental visit, mammogram, and prostate test. For cholesterol test and dental visit, significant changes can be observed starting 6 years prior to dementia. The absolute difference increases to 5 percentage points about 2 years prior to the incidence and reaches 8-10 percentage points by the time of dementia incidence.

Significant reductions are also found in physical activities and social engagement, as manifested in Figure 6. The impact emerges approximately 2-4 years prior to the incidence of dementia and continues to increase over time. Particularly, we find that moderate and light physical activities decline about 8 percentage points among the dementia sample relative to the control sample 2 years prior to the incidence of dementia. Furthermore, the incidence of dementia is associated with about 15 percentage points declines in moderate and light activities, indicating a substantial impact; and the impact tends to persist after the incidence of dementia.

## 5. Robustness Analysis and Placebo Test

### 5.1 Sample Selection and Model Specifications

In the main analysis, we use 1:1 propensity score matching (PSM) without replacement to form balanced pairs of dementia and control samples. However, this approach may be overly restrictive, excluding many potential control samples. To address this concern and refine the estimation of common trends, we conducted robustness analysis using two alternative approaches: 1:2 PSM with replacement (Austin, 2010) and inverse probability weighting (IPTW) (Chesnaye et al., 2022). The results, presented in Figure A1-A3 (others available upon request), show that the findings are robust to these alternative sample selection and model specifications.

Another concern is the age of the study sample. While we do not impose special age restrictions in the main analysis to fully observe the periods before the incidence of dementia, the changes in Medicare enrollment over time, which typically begins at age 65, could potentially affect the estimation of dynamic effects, particularly for preventive care. Therefore, in the robustness analysis, we further restrict the sample to individuals aged 65 and older. The results, shown in Figure A1-A3, demonstrate the robustness of the findings to the age restriction.

### 5.2 Aging and Cohort Effects

The event study estimates depend on the model of common trends. In the main analysis, we include age to adjust for aging trends, and year fixed effects and year-by-region fixed effects to account for time trends. However, the specification may not fully capture the aging and cohort effects. To address this concern, we conduct a series of robustness analyses by more flexibly controlling for these effects. Specifically, we (a) include age and age squared in the estimation, (b) use 5-year age bins, (c) introduce 1-year age bins (age fixed effects), and (d) include both 1-year age bins and birth year fixed effects. The results, available upon request, are very similar to our main estimates, indicating the robustness of our findings.

### 5.3 Placebo Tests

Lastly, we perform placebo tests to demonstrate that the observed patterns are unique to dementia and not applicable to other health shocks. Specifically, we repeat our analysis by estimating the change in behavioral outcomes before and after the incidence of a particular chronic or acute condition relative to the matched controls who never develop such condition. Two common chronic conditions and two common acute conditions are examined as negative controls, including diabetes, hypertension, hip fracture, and myocardial infarction^1^ (Nicholas et al., 2021). The patterns of these conditions differ significantly from those of dementia as shown in Figures A4-A6.^2^ Specifically, there are no significant reductions in preventive care utilization and healthy behaviors associated with these conditions, especially during the periods before the incidence. In some cases, such as diabetes and hypertension, the incidence is even associated with increased utilization of preventive care. These findings support the uniqueness of the patterns observed for dementia and emphasize the early presentation of this disorder.

## 6. Conclusion and Discussion

Using nationally representative longitudinal data in the US, this study provides important insights into the long-term impacts of dementia on preventive care utilization and health behaviors. Our findings reveal significant and considerable declines in preventive care utilization and healthy behaviors among individuals affected by dementia. Notably, these declines manifest years prior to the actual onset of dementia, intensify over time, and persist for years following the diagnosis. This pattern underscores the vulnerabilities and unmet necessities in managing health within this population.

Importantly, while there is a noticeable rise in memory-related diagnoses around the time of dementia incidence, a pronounced underdiagnosis issue becomes evident, as being recognized in the literature (Alzheimer’s Association, 2023). A substantial proportion of individuals with dementia experience late diagnoses or even remain unaware of their condition, thus accentuating the long-term negative implications of dementia on health-related choices and behaviors, as well as medial and societal costs (Getsios et al., 2012).

The implications of the declines in preventive care utilization and healthy behaviors among dementia patients are profound. Missed cholesterol tests and dental visits, for example, can increase the risk of cardiovascular diseases and dementia, leading to a higher disease burden and healthcare costs (Fereshtehnejad et al., 2018; Livingston et al., 2020; Mortensen and Nordestgaard, 2020; Sud et al., 2020; Winblad et al., 2016). Recent evidence also suggests that dementia is associated with increased emergency department visits, hospitalizations, and medical costs, even before a formal diagnosis (Desai et al., 2019; Lin et al., 2016). Hence, our findings highlight the substantial healthcare burden and costs that may arise due to disrupted preventive care and health behaviors in the context of dementia.

Detecting early signals of dementia is thus pivotal as it may provide several key benefits. First, it allows individuals and families to better prepare for the future and make informed decisions about their care. Second, it prompts individuals and families to seek appropriate services and consultations, shielding them from potential adverse behavioral and health consequences. As early signals of dementia manifest consistently across preventive care and behavioral health contexts long before a diagnosis, collaboration among primary care providers, caregivers, and other stakeholders is crucial to efficiently identify and act upon these signals for effective planning and support (Bannenberg et al., 2021). Moreover, early signals of dementia have also been found in the financial sector, transportation sector, etc (Bayat et al., 2021; Eyigoz et al., 2020; Nicholas et al., 2021). Harnessing precursors of dementia within the health sector, such as through preventive care and behavioral health in our context, serves as a pilot towards challenging but potentially more fruitful cross-sector collaborations in early detection of dementia.

Several limitations should be acknowledged. First, our reliance on self-reported measures may introduce recall bias. Future studies could consider linking Medicare claims data to the HRS to obtain more accurate measures of healthcare utilization. Second, although we accounted for time trends as well as aging and cohort effects, changes in clinical guidelines and their applicability to different population groups for certain preventive care measures could still influence our estimation and interpretation. Third, the use of unbalanced panel data may affect the accuracy of estimates for time periods further away from the incidence of dementia. Future studies with larger samples can help address this issue. Lastly, the mechanisms underlying the observed declines were not examined in this study. Factors such as cognitive barriers, stigma, distorted risk perception, and lack of caregiving may contribute to these behavioral changes and warrant further investigation.

In conclusion, our study demonstrates that dementia is associated with reduced preventive care utilization, physical activities, and social engagement years before its onset, with these effects persisting and escalating after the incidence of dementia. These findings underscore the significant negative consequences of dementia on health-related decision-making and well-being. To target preventive measures more effectively, we emphasize the need for additional and consistent early signals of dementia and cross-sector cooperation. By identifying better strategies and interventions for those most likely to benefit, the overall management and outcomes can be improved for individuals living with dementia.

## Data Availability

All data produced in the present study are available upon reasonable request to the authors.

## Acknowledgements

This study was funded by a research grant R01AG077529 from the U.S. National Institute on Aging; career development award K01AG053408 from the U.S. National Institute on Aging; and grant P30AG021342 from the U.S. National Institute on Aging to the Yale Claude D. Pepper Older Americans Independence Center. The funders had no role in the study design; data collection, analysis, or interpretation; in the writing of the report; or in the decision to submit the article for publication.

## Figures

**Figure A1.**
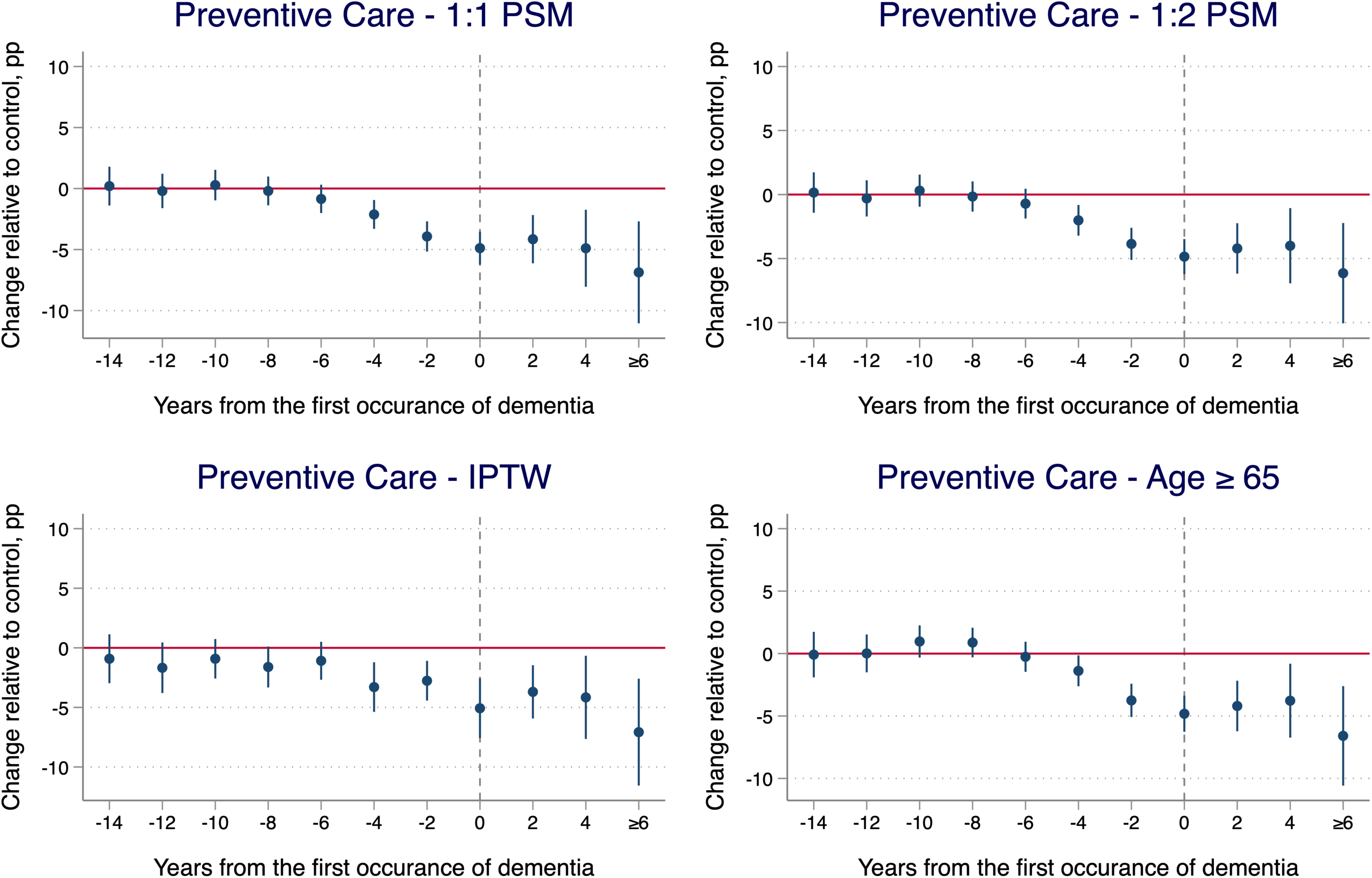
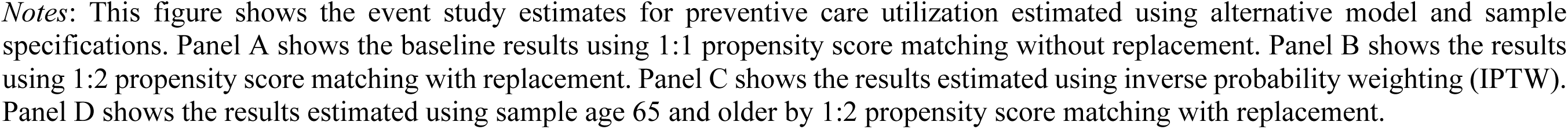
Robustness of the event study estimates for preventive care utilization to alternative model and sample specifications.

**Figure A2.**
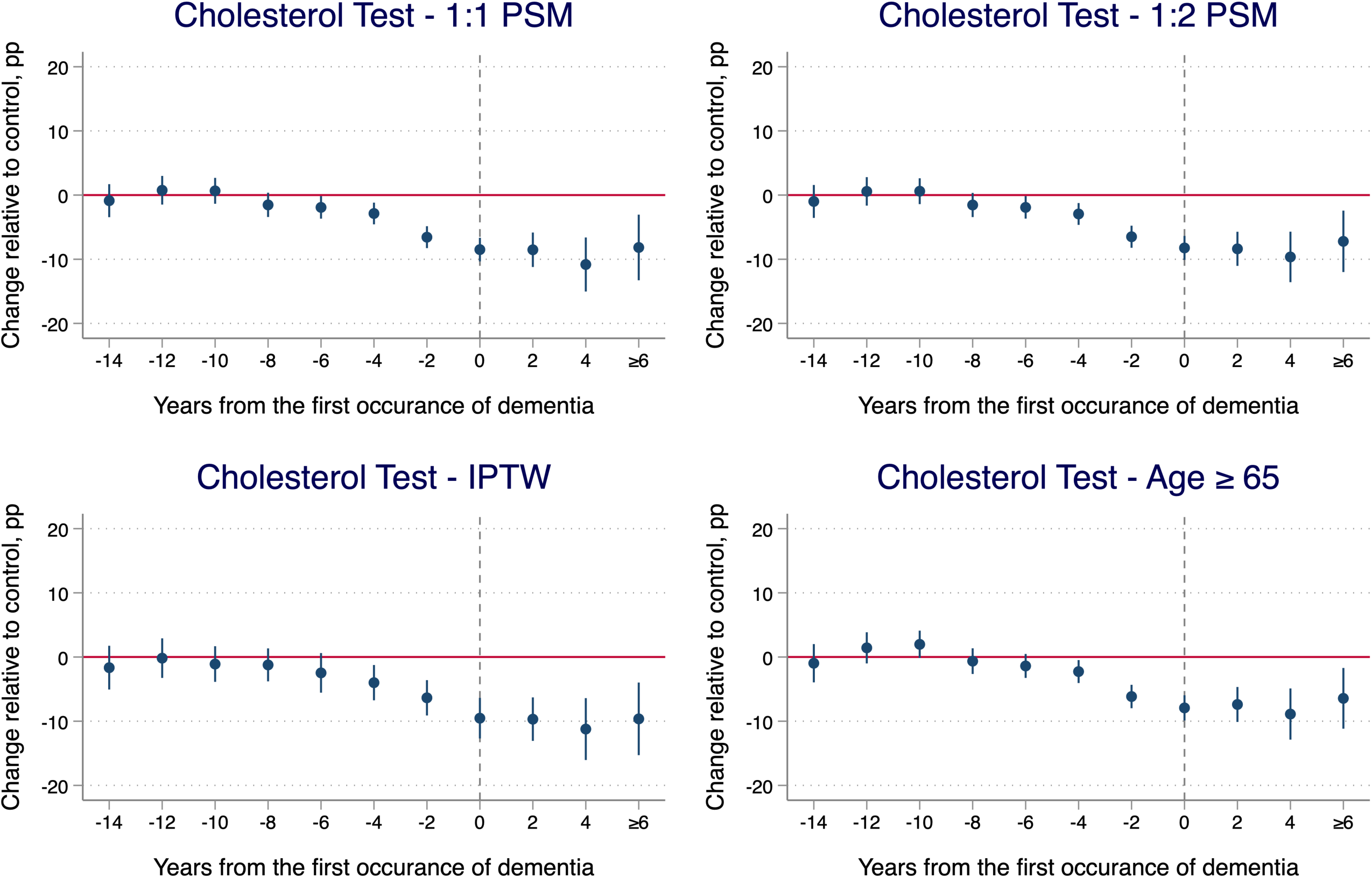
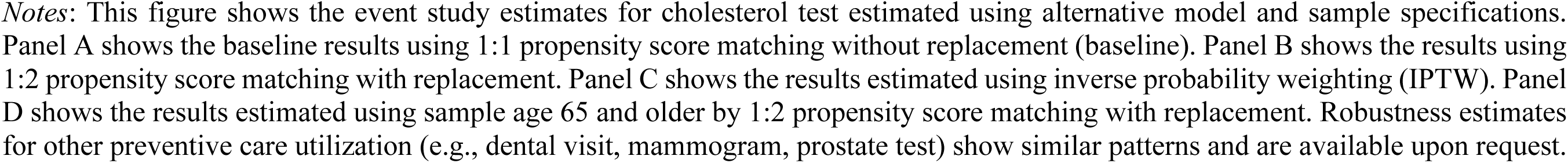
Robustness of the event study estimates for cholesterol test to alternative model and sample specifications.

**Figure A3.**
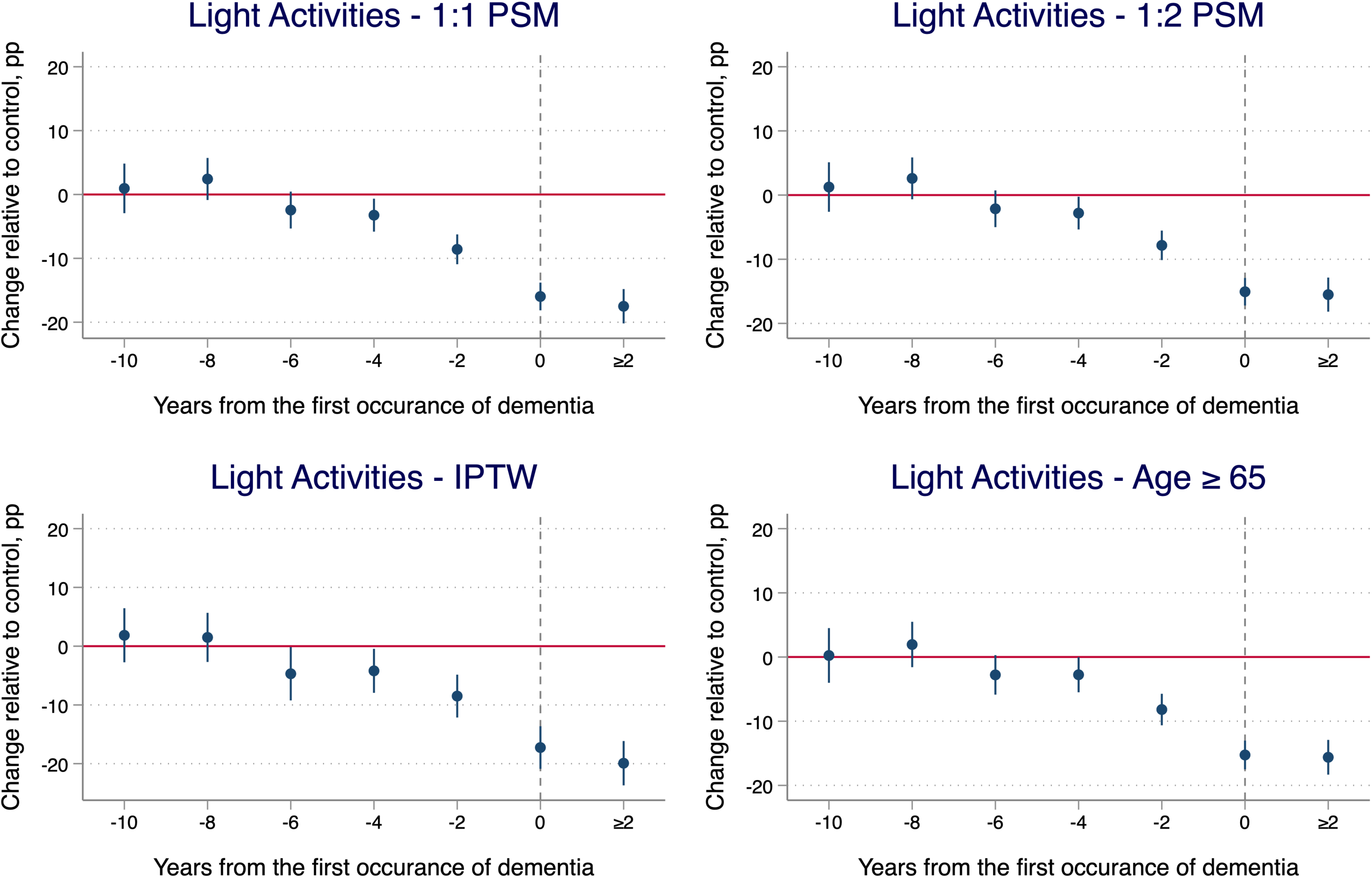
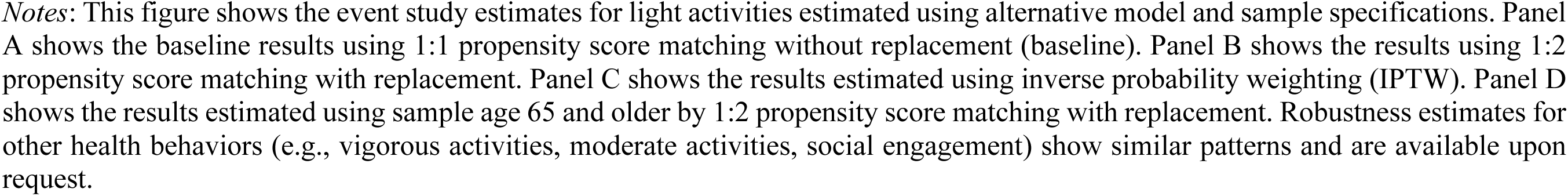
Robustness of the event study estimates for physical activities to alternative model and sample specifications.

**Figure A4.**
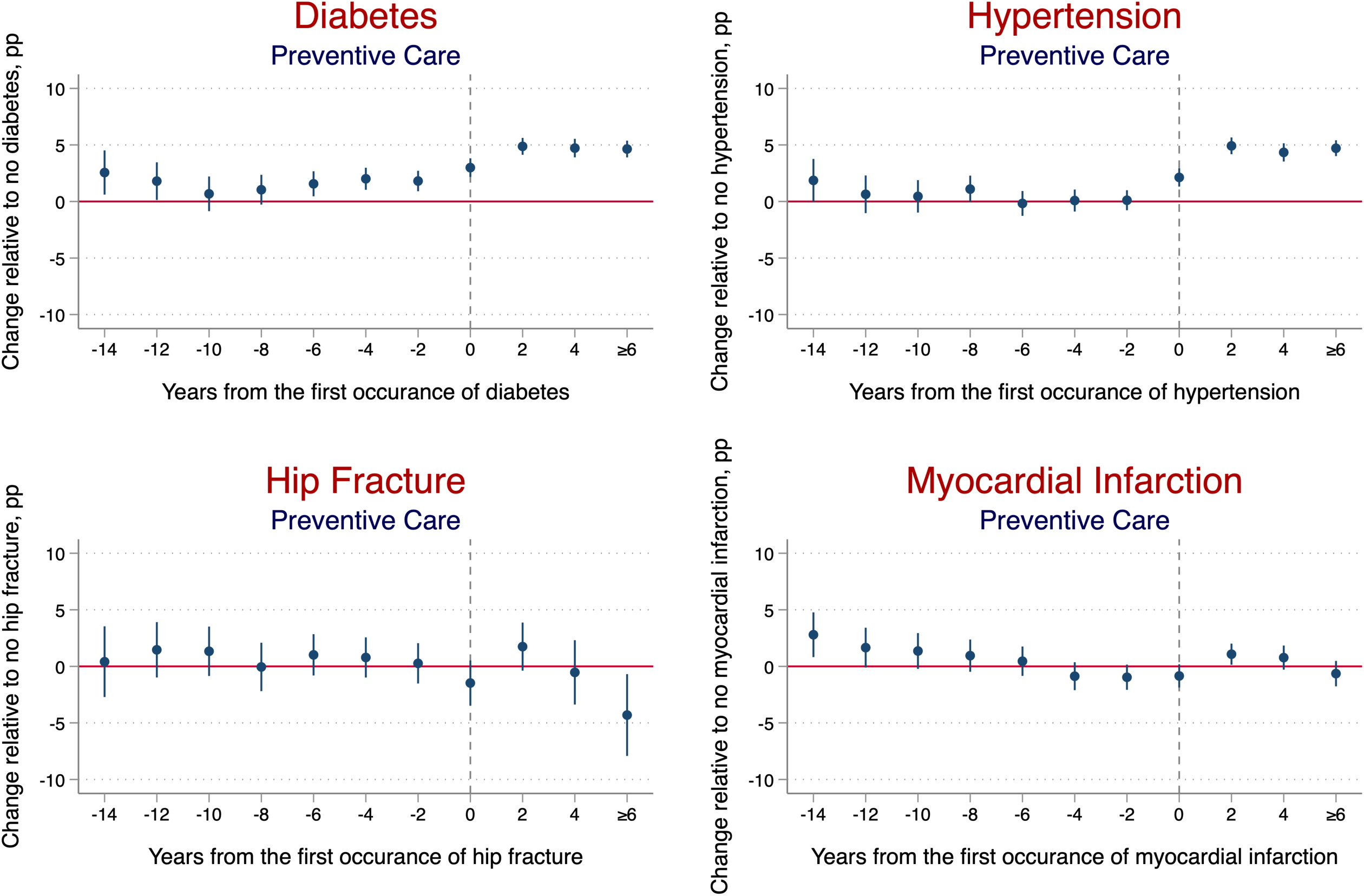
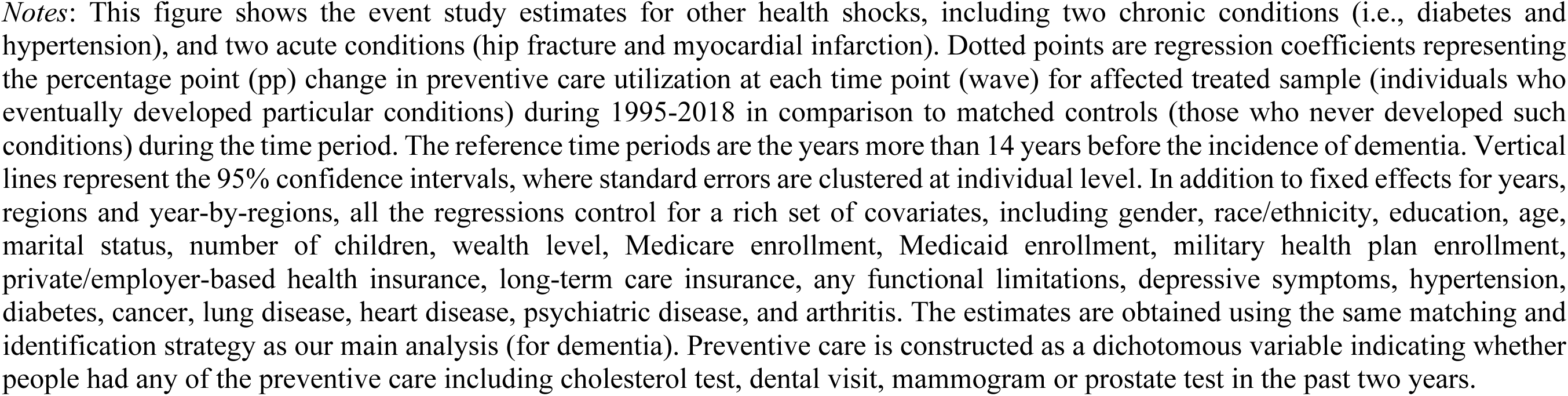
Placebo test: change in the preventive care utilization before and after the incidence of other chronic or acute conditions for affected treatment sample relative to matched control sample during 1995-2018.

**Figure A5.**
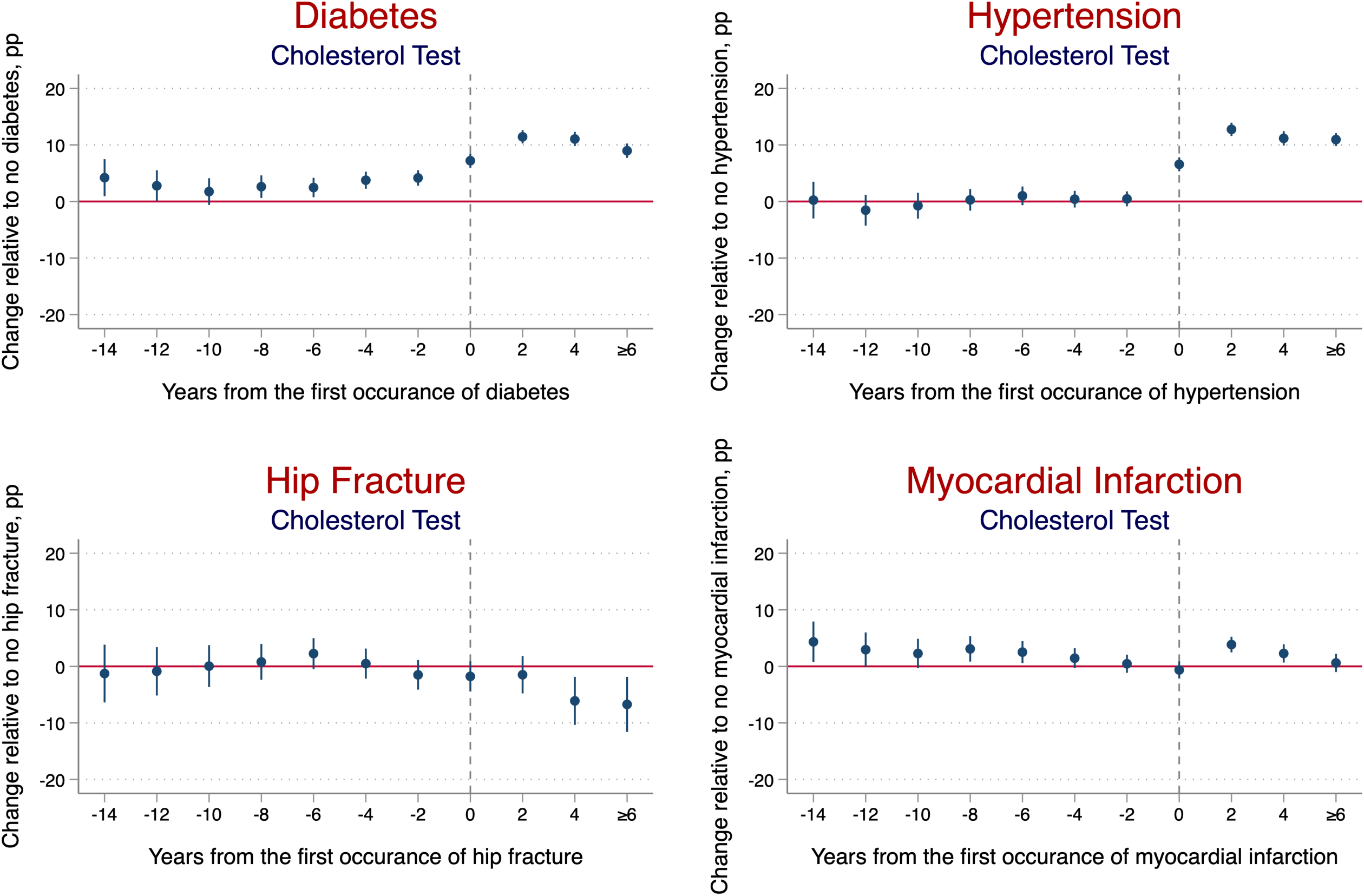
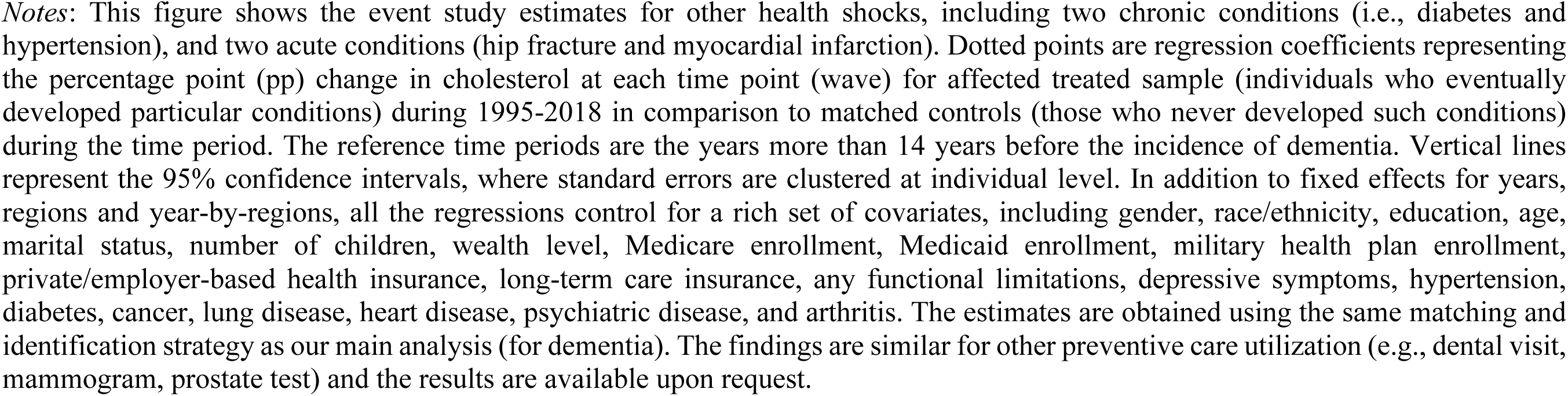
Placebo test: change in the cholesterol test before and after the incidence of other chronic or acute conditions for affected treatment sample relative to matched control sample during 1995-2018.

**Figure A6.**
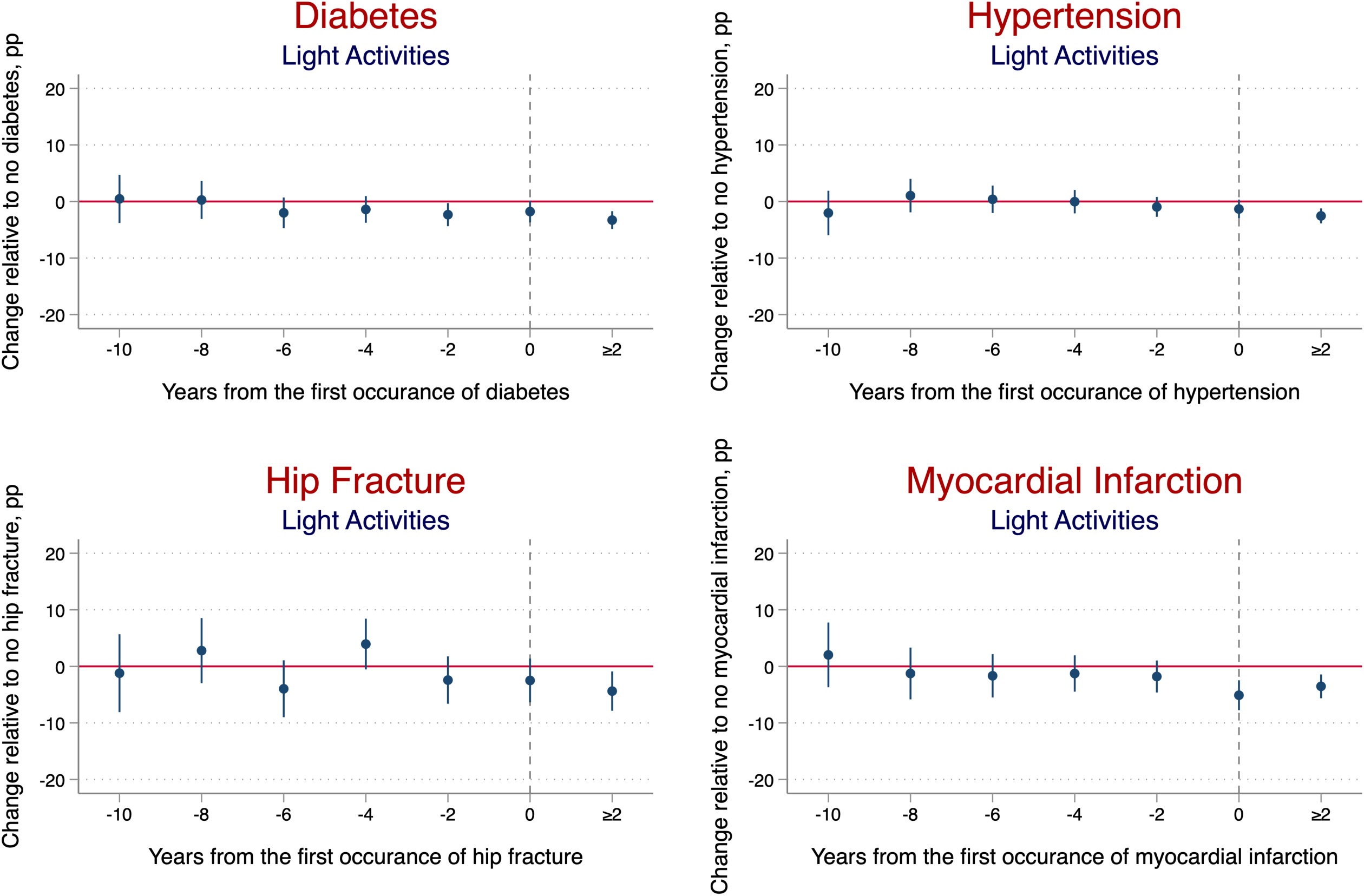
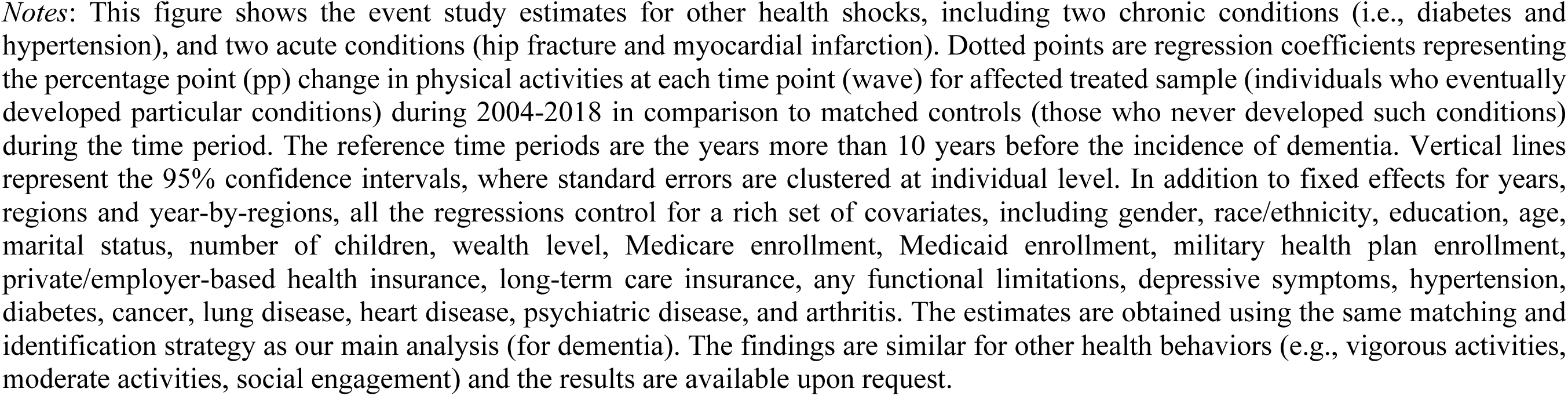
Placebo test: change in the physical activities before and after the incidence of other chronic or acute conditions for affected treatment sample relative to matched control sample during 2004-2018.

## Footnotes

1 Some other conditions, such as arthritis and cancer, are also examined. The findings of these conditions are very similar to the ones included.

2 The results for other outcomes are available upon request. The findings are similar.

